# Pre-existing antibodies and age shape the immune response following EV-A71 vaccine: a prospective serological study among the Chinese pediatric population

**DOI:** 10.1101/2025.09.17.25335968

**Authors:** Jiaxin Zhou, Xiaoyu Zhou, Junbo Chen, Nicholas C Grassly, Hongjie Yu

**Author notes:** These two authors contributed equally to this work (Jiaxin Zhou, Xiaoyu Zhou). **Correspondence:** Prof. Hongjie Yu, School of Public Health, Fudan University, Key Laboratory of Public Health Safety, Ministry of Education, Shanghai 200032, China.,.

## Abstract

Hand, foot, and mouth disease (HFMD), primarily caused by enterovirus A71 (EV-A71), poses significant pediatric health risks in the Asia-Pacific region, with vaccination being critical for prevention. Using longitudinal serological data from 1 585 Chinese children in a 2019–2020 prospective study (combining clinical trial and cohort samples), the present study assessed how pre-vaccination antibody titers and age influence EV-A71 vaccine response. EV-A71-specific antibodies were measured via a modified cytopathogenic effect assay, with geometric mean titers (GMTs) and fold increases (GMFIs) analyzed. At baseline, 3.9% (N = 61) of children were seropositive. After vaccination, those with high pre-existing titers exhibited higher GMTs, compared to those with low-level or undetectable titers (GMTs: 10.30 [95% CI: 9.09, 11.52] *vs*. 7.58 [95% CI: 6.86, 8.29] *vs*. 6.92 [95% CI: 6.82, 7.02]; GMFIs: 3.36 [95% CI: 1.97, 4.75] *vs*. 4.23 [95% CI: 3.48, 4.98] *vs*. 4.92 [95% CI: 4.82, 5.02]).

Generalized additive models revealed that predicted GMTs peaked (8.78) at a baseline titer of 10.20, while GMFIs declined with higher pre-vaccination titers. Vaccine-induced responses also varied with age, showing periodic increases in children under 36 months. Collectively, these findings quantify the effects of pre-existing antibodies and age on the antibody level following vaccination, highlighting the importance of tailoring immunization schedules based on both pre-existing immunity and age. Accordingly, immune-naïve children should be vaccinated as early as possible.

## INTRODUCTION

Hand, foot, and mouth disease (HFMD) is a common childhood disease primarily affecting children under 5 years old, caused by multiple serotypes of enterovirus. It imposes a high disease burden in the Asia-Pacific region, especially in mainland China, where more than 15 million HFMD cases were reported between 2013 and 2019, including 77 000 severe cases^1^. Enterovirus A71 (EV-A71) and Coxsackievirus A16 (CVA16) are the major pathogens, with EV-A71 related to around 70% severe cases and 90% deaths associated with HFMD^1–3^, posing a significant threaten to pediatric health. Given the lack of a specific antiviral drug for HFMD treatment, vaccination remains the most effective way to prevent and control the disease. In 2016, three types of monovalent inactivated EV-A71 vaccines were licensed in mainland China, with high vaccine efficacy over 90% for children aged 6 months to 5 years^4,5^. Although the proportion of severe cases and mortality has decreased since 2016, CVA16 and other coxsackieviruses, such as CVA6, are becoming the dominant serotypes. This shift highlights the urgent need for a bivalent EV-A71-CVA16 vaccine and other multivalent vaccines to broaden protective coverage.

Vaccine-induced antibodies are crucial for protecting vulnerable neonates and younger children after the decline of maternal antibodies, and heterogeneity of immune response post-vaccination has been associated with host factors and vaccines^6^. Previous research on various infectious diseases (e.g., measles, influenza, hepatitis B, poliovirus, pertussis, and tetanus) has revealed that pre-existing antibodies (including maternal, infection-induced and vaccine-induced antibodies) can interfere with both humoral and cellular immunity responses following vaccination^7–10^, posing challenges to vaccine development and strategy. However, for EV-A71, most previous studies have only reported on the immunogenicity of the EV-A71 vaccine across different age groups, without clarifying the baseline titers of participants or focusing primarily on children with undetectable pre-vaccine titers^11,12^. The potential effects of pre-vaccine antibodies and age at vaccination on immune response post EV-A71 vaccination remain largely unclear, warranting further research.

A more comprehensive survey of previously unanalyzed data from the complete series of monovalent EV-A71 vaccine clinical trials presents a unique opportunity to investigate the influence of pre-existing antibodies and other potential factors on immune response with a larger sample size. Therefore, based on the combined dataset from a clinical trial and a cohort study among the Chinese pediatric population under 4 years of age, our study aimed to investigate the effects of pre-existing antibody titers and vaccination age on vaccine-induced immune responses in children with varying pre-vaccine titer levels, after adjusting for multivariate confounders.

## RESULTS

A total of 1 585 children were included in the analysis from the Phase 3 clinical trial (N=638) and the observational study (N=947). The demographic characteristics of participants are displayed in **Table 1**. Among children included, 789 (49.8%) were girls and the median age at baseline was 17.5 months (IQR: 11.3-26.9 months). The proportion of boys among children from the clinical trial was lower than that of children from the cohort study (***p*** = 0.0322). Besides, a significantly greater number of children aged 6-11 months were enrolled in the clinical trial (***p*** < 0.0001). A comparison of baseline characteristics between children enrolled and not enrolled in this study is presented in **Supplementary Information Note 4**.

**Table 1.**
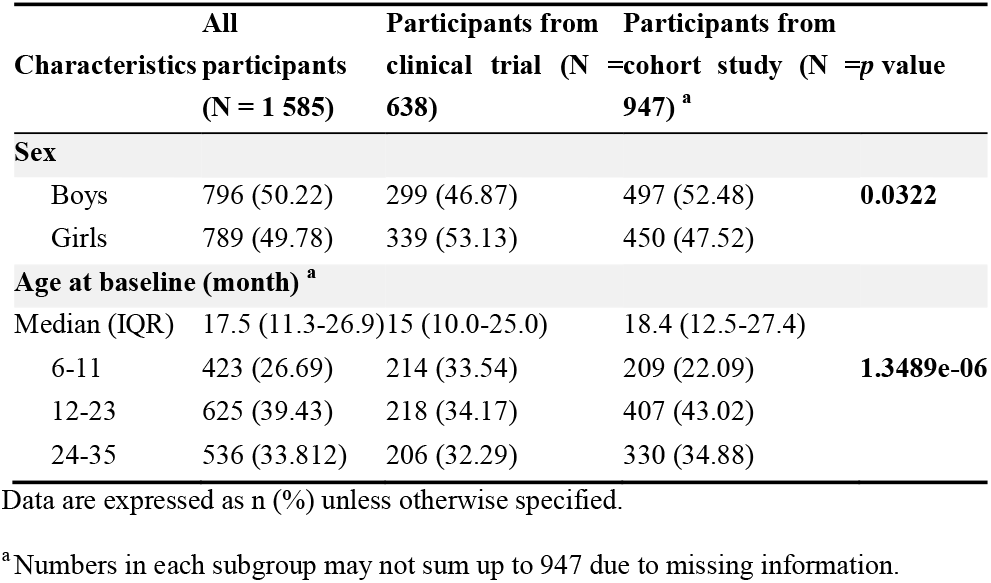
Characteristics of children enrolled in this study.

According to the baseline titer, the majority of participants were seronegative, with only 3.9% (n=61/1 585) being seropositive, including 24 (1.5%) and 37 (2.4%) children with high- and low-levels of pre-existing antibodies, respectively. Most individuals showed a significant increase in antibody titer from baseline to 30 days post 2-dose vaccination (**Figure 1A, Supplementary Information Note 5**), and the overall GMT following two doses of vaccination was 6.99 (95% CI: 6.89, 7.09). A clear pattern was observed, with higher levels of GMT induced by 2 doses of vaccination observed among children with higher levels of pre-existing antibody, compared with those with low-level or undetectable baseline titers (10.30 [95% CI: 9.09, 11.52] *vs* 7.58 [95% CI: 6.86, 8.29] *vs* 6.92 [95% CI: 6.82, 7.02]; ***p*** = 7.8567e-08) (**Figure 1B**). However, a reverse trend was observed regarding the fold increases of antibody titer following 2 doses of vaccination. The GMFI of children with higher pre-existing titers was 79.4% and 68.3% of that in children with low or undetectable baseline titers (3.36 [95% CI: 1.97, 4.75] *vs*. 4.23 [95% CI: 3.48, 4.98] *vs*. 4.92 [95% CI: 4.82, 5.02]; ***p*** = 0.0191) (**Figure 1C**). We further analyzed the vaccine-induced titers and fold increases across doses, observing similar results to those in the primary analysis (**Supplementary Information Note 6.1**).

**Figure 1.**
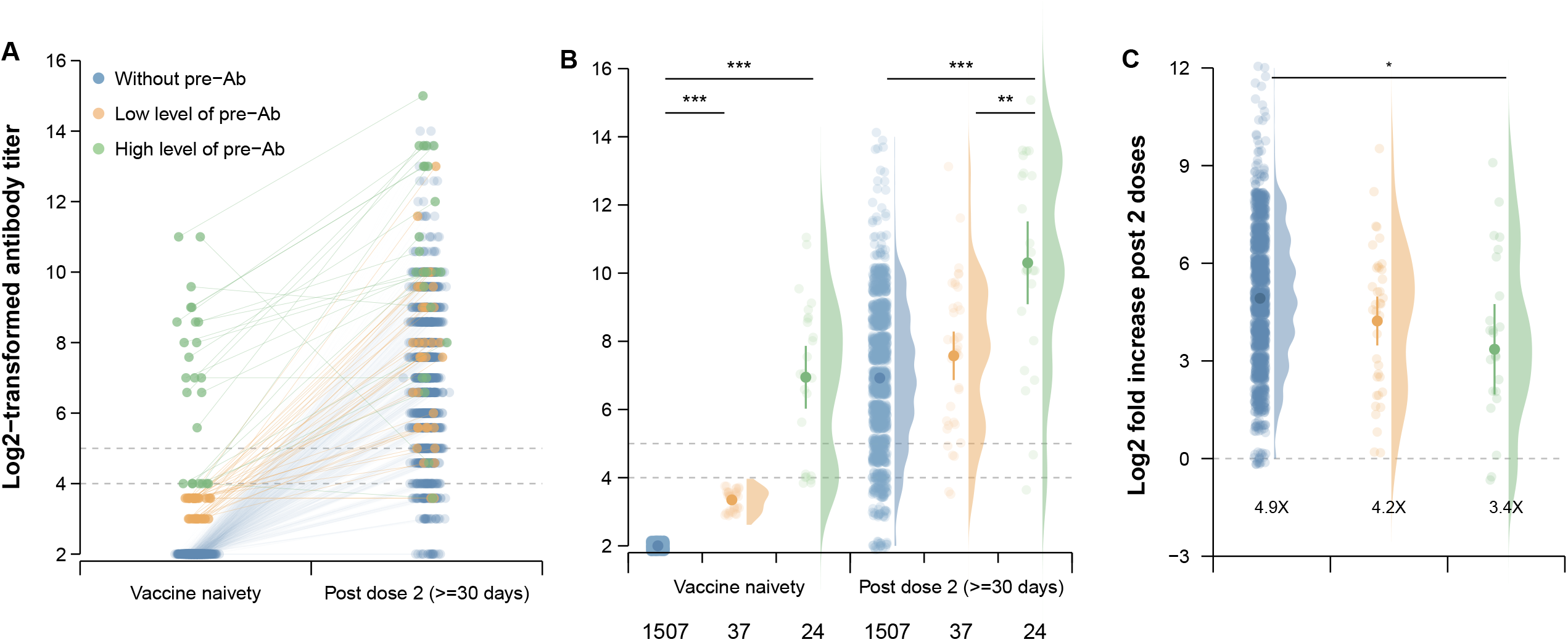
Immune response to the EV-A71 vaccine from vaccine naivety to 30 days post dose 2, by the level of pre-existing antibody. Panel (A) shows the individual antibody trajectories from vaccine naïvety to 30 days after receiving two doses of vaccination. Panel (B) shows the distribution and GMTs of antibodies from vaccine naïvety to 30 days after receiving two doses of vaccination. Panel (C) shows the log2-transformed fold increase of antibody titers after 2 doses of vaccination, and the geometric means of fold increase are marked at the bottom. Note that the figures in panel (B) below show the sample size of each group. “*”, “**” and “***” represent the *p*-value of the statistical test is <0.05, <0.01 and <0.001. The dashed grey lines in panels (A) and (B) denote the selected seropositive thresholds (i.e., log-transformed titers of 4 and 5).

Figure 2. illustrates the predicted average titers and the fold increase post-vaccination over pre-existing titers, after controlling for sex, vaccination age at the primary dose, date of sampling, and source of data. With the increase in pre-existing antibodies, the vaccine-induced titers increased from 5.70 when the baseline titer was 2.00, reaching their peak of 8.78 (95% CI: 7.32, 10.23) when the baseline titer was 10.20, followed by relative stability (**Figure 2A**). Overall, the fitted titer after vaccination consistently exceeded the seroprotective thresholds irrespective of pre-vaccination titers (**Figure 2A**). On the contrary, the fold increases in antibody titer exhibited an inverse pattern and declined almost linearly, with an initial level of 3.86-fold when baseline titer was 2.00 and reached to its lowest value of 0.78-fold (95% CI: −0.79, 2.35), where the corresponding titer was 12.00 (***p*** = 0.0004; **Figure 2B**). The results of the sensitivity analysis were not significantly different from those of the primary analysis (**Supplementary Information Notes 6.2 and 6.3**).

After assessing the potential effect of age at the first dose, we observed similar patterns among children with varying baseline titers. Between 6 and 36 months of age, the vaccine-induced antibody titers increased periodically, reaching their first highest and lowest points when the first dose was administered at 10.20 and 19.12 months of age, respectively (**Figure 3**). The mean curve of predicted vaccine-induced titers exceeded the predefined thresholds irrespective of the vaccination age at the first dose. Although the curves of prediction were almost parallel, the predicted titers following two doses of vaccine were higher among children with high-level pre-existing antibodies compared with their counterparts with low levels of pre-existing antibodies (***p*** = 0.0016; **Figure 3A-B**). The predicted titers among children in the negative group were lower than children from the positive group and remained close to the seroprotective thresholds (**Figure 3C**).

**Figure 2.**
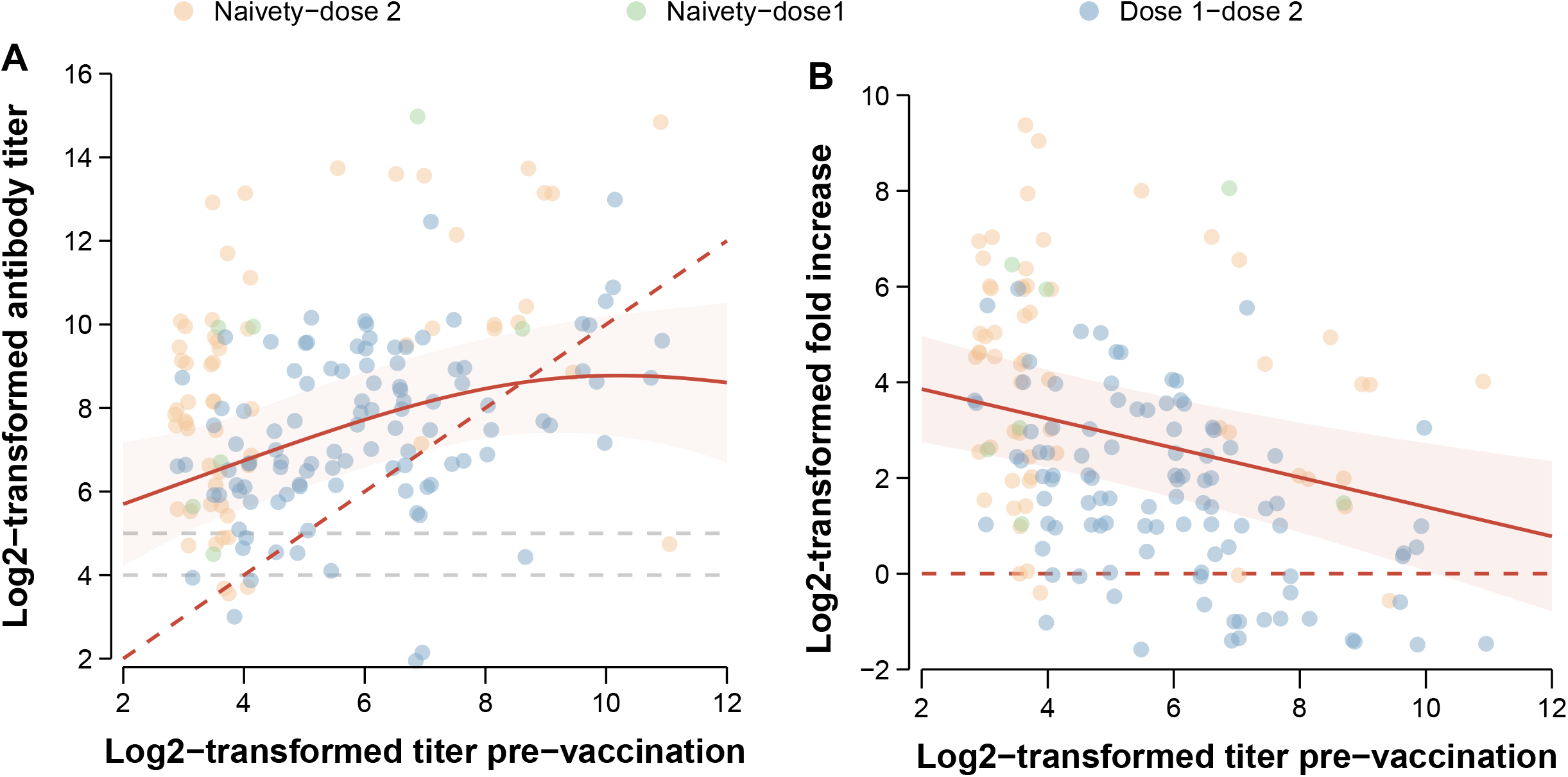
Fitted vaccine-induced antibody titers and fold increase using GAMMs. Panel (A) shows the fitted post-vaccination antibody titers by the levels of pre-existing titer. Panel (B) shows the fitted fold increase post-vaccination by the levels of pre-existing titer. Note that the solid lines and shadows indicate the fitted mean and 95% CI band. The red dashed lines in the two panels indicate the reference line. The grey dashed lines denote the selected seropositive thresholds (i.e., log-transformed titers of 4 and 5).

**Figure 3.**
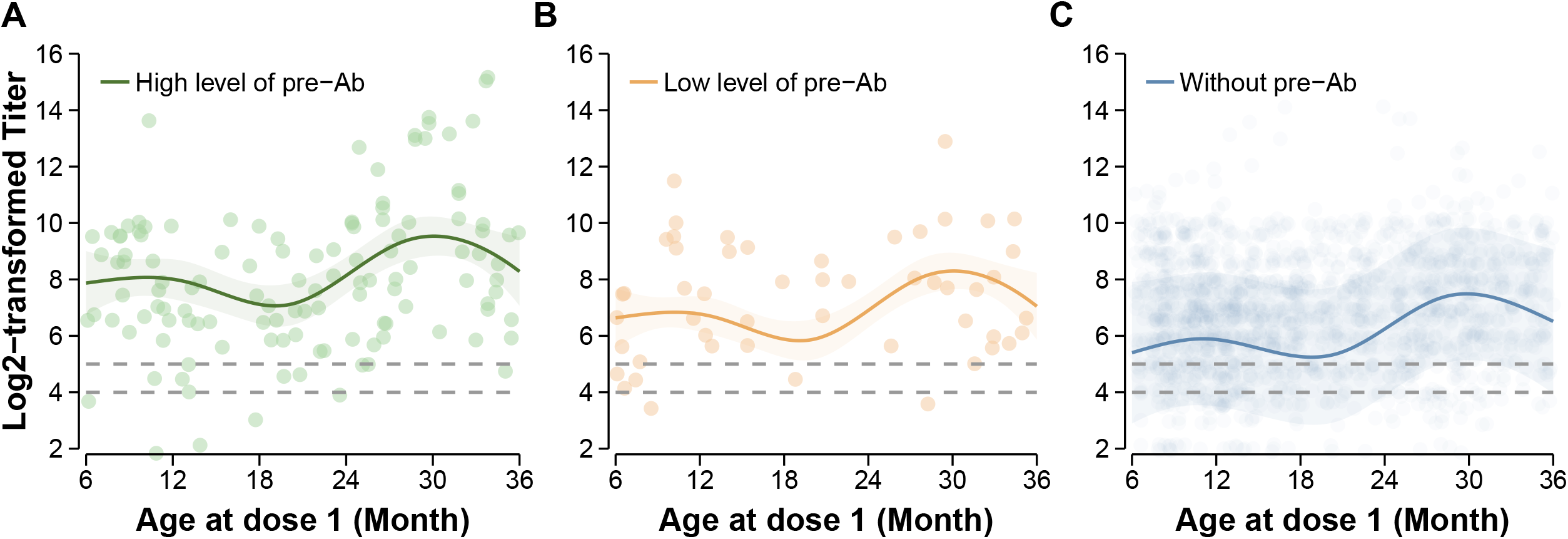
Effect of vaccination age at first dose on the immune response post-dose 2, stratified by pre-existing antibody levels. Panels (A) - (C) show the age effect among children with high- and low-levels of pre-existing antibody and children without pre-existing antibody, respectively. The points represent individual log-transformed antibody titers following 2 doses. The lines are predicted curves of post-vaccination antibody titers. The shadow areas indicate the 95% CIs of the predicted curves. The dashed grey lines denote the selected seropositive thresholds (i.e., log-transformed titers of 4 and 5).

## DISCUSSION

In this study, we demonstrated the significant impact of pre-existing EV-A71 antibody levels and age at primary vaccination on the immune response to EV-A71 vaccination among younger Chinese children. Our findings highlight that a higher level of pre-existing antibodies against EV-A71 prior to vaccination is inversely associated with the increase in the fold of antibody response after vaccination with an inactivated EV-A71 vaccine. Nevertheless, children with higher pre-existing antibody levels still achieved higher post-vaccine antibody titers overall. Besides, we observed a periodic increase in the vaccine-induced antibody titers with age at vaccination, after adjusting for other factors, including pre-existing antibody levels. These findings suggest that pre-existing antibody levels may dampen the apparent antibody boosting by the EV-A71 vaccine and age has a positive influence on shaping the immune response to EV-A71 vaccination. These insights are crucial for the development of enterovirus vaccines and underscore the importance of tailoring vaccination strategies to both pre-existing immunity and age to administer the primary dose.

We found that the overall GMT following 2 doses of vaccination exceeded the pre-defined seroprotective thresholds and indicating robust immune protection regardless of the pre-vaccination titers, consistent with the previous results from a clinical trial (GMT: 142.7, 95% CI: 128.8, 158.2)^4^. The estimated GMT and fold increases were lower than those reported by Zhu et al.^5^ (GMT: 325.3, 95% CI: 284.8, 371.7), which might be attributed to the lower proportion of children with pre-existing immunity in our study (3.8% *vs*. 27.2%). These potential limiting effects of pre-existing immunity on immunity boosting by vaccination were consistent with previous serological studies of other infectious diseases, such as measles and influenza^7,13–15^. In our study, the pre-existing antibody titre may stem from prior asymptomatic EV-A71 infection or cross-activity by asymptomatic/mild-clinical infection of other serotypes or extremely limited existing maternal antibody, which might mask the presentation of viral epitopes of antigens administered intramuscularly to form complexes, thereby attenuating the stimulation of naïve or memory B cells^9,15^. It’s also important to note that the observed inverse association between pre-existing antibody level and fold change could alternatively be explained by a physiological ceiling effect or the limitation of serological assay, which may constrain the accurate measurement of higher antibody titers.

Moreover, we observed a positive trend in antibody titers after full-schedule vaccination, as the age at the first dose increased, which has also been reported for other vaccines, including oral polio vaccine (OPV)^16^, diphtheria-tetanus-pertussis (DTP) vaccine^17^, and measles vaccine^18^. The periodical increase in vaccine-induced immunity may be associated with a combination of factors, including the attenuation of the pre-vaccination antibodies, the development of the immune system^7^, and the increasing rate of asymptomatic infection from other serotypes and genotypes of enteroviruses with age^19^. These results, together with the effect of baseline titers we have mentioned, suggest that optimal vaccination strategies should consider both the levels of pre-existing immunity and children’s ages. Our results suggest that young children, especially those who are immune-naïve due to the elimination of maternal antibodies after 2 months of age^20^ or those without infections, should be vaccinated as early as possible. Besides, for the development and clinical trials of HFMD vaccination, it is essential to consider the pre-existing antibody titer and age distribution in the population to achieve optimal vaccine effectiveness.

It has been reported that pre-existing antibodies and age play a critical role in shaping the dynamics of vaccine-induced immunity, influencing not only response levels but also the patterns of antibody decay^14,21^. Thus, among children with varying baseline titers and ages, heterogeneity in the long-term dynamics of antibodies following EV-A71 vaccination may exist, which is relevant to the duration of immune protection provided by the EV-A71 vaccine. However, due to data limitations, this study focused solely on the short-term responses following vaccination and was unable to address that evidence gap. Therefore, longitudinal serological cohort studies are needed to further evaluate and compare the differences in the durations of vaccine-induced immunity and susceptibility to EV-A71 infection among children with various immune backgrounds, which would be quite helpful in identifying high-risk populations and implementing strategies to bridge the immunity gaps. Moreover, the cellular immunity post-vaccination, as an essential part of the immune response, could not be evaluated in this study. Future studies could investigate the quantified effect of pre-existing immunity on vaccine-induced cellular immunity. In addition, we only measured the EV-A71-specific antibodies in this study; the effect of cross-reactivity by other serotypes on the EV-A71 vaccine immunity was not available to evaluated in this study and needs further analysis in the future, although previous studies supported that a monovalent inactivated EV-A71 vaccine provided limited cross-protection against HFMD associated with other serotypes^22,23^.

In conclusion, this study provides hitherto undocumented evidence of the effect of pre-existing antibodies and age at the time of the primary dose on the immune response to the EV-A71 vaccine. Our findings suggest that pre-existing antibody levels and age play essential roles in tailoring immunization strategies to achieve better responses to the EV-A71 vaccine. Especially for children with no prior exposure to enteroviruses, vaccination should be conducted as early as possible. Besides, long-term serological cohort studies based on the general population are essential for further evaluating the dynamics and long-term vaccine-induced immunity among children with diverse immune backgrounds, particularly during early life.

## MATERIALS AND METHODS

### Study design and participants

Data on children’s humoral immune response before and after EV-A71 vaccination were derived from two serological studies conducted in Southern China between 2019 and 2020: (a) a phase 3 clinical trial conducted to test the equivalence of the immunity responses of three batches of inactivated EV-A71 vaccines among healthy children aged 6-35 months old between September 2019 and April 2020 in Sichuan Province (NCT07026500); (b) a cohort study carried out in Hubei Province between April 2019 and March 2020 to assess the immune response following EV-A71 vaccination and estimate the vaccine effectiveness (Jiaxin Z, 2025, unpubl. data). Children with known HFMD history or those who have already received the EV-A71 vaccine were excluded at enrollment for both studies. Participants with paired serum samples before the first vaccination dose and 30 days after the second dose were included in this study. Moreover, a subset of children selected from a clinical trial by age stratification was sampled at 7, 14, or 30 days after the first dose and at 7, 14 days after the second dose (**Figure 4, Figure S1**). The details of the study design are presented in **Supplementary Information Note 1**. In both studies, participant epidemiological information (i.e., sex, date of birth, and ethnicity) was collected at baseline, and the dates of vaccination and sample collection were accurately recorded. Information and data were recorded, corrected, and checked using an electronic case report form to ensure data security. Children were included in this study when they: 1) received 2 doses of EV-A71 vaccine after recruitment; 2) provided samples before and after vaccination; 3) had provided accurate data of demographic information and dates for vaccination (**Figure 4**). The detailed sampling time point and corresponding sample sizes are depicted in **Figure S1**.

**Figure 4.**
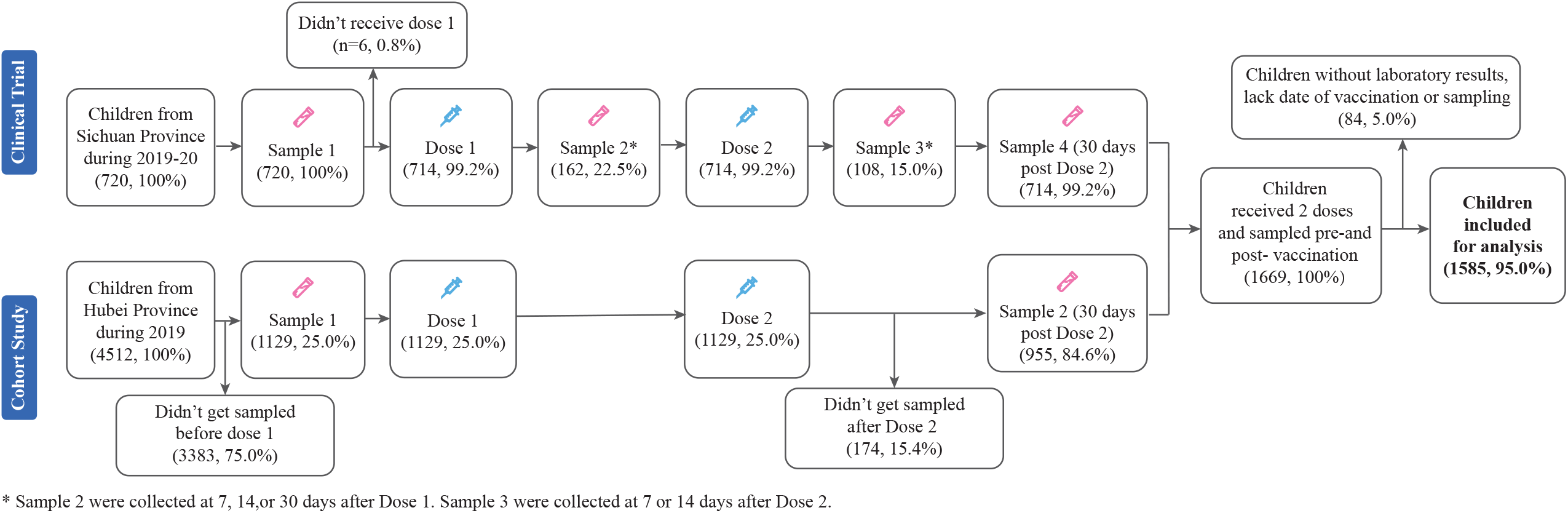
Overview of the study design. Participants in both the observational study and the clinical trial provided paired samples before vaccination and 30 days after completion of EV-A71 vaccination. In addition, a subgroup of participants from the clinical trial was selected (stratified by age group) and provided Sample 3 or Sample 4.

This study was approved by the Institutional Review Board of the Sichuan Provincial Centre for Disease Control and Prevention (No. SC-0820190301) and Hubei Provincial Centre for Disease Control and Prevention (No. 2019-001-02). Written informed consents of all enrolled children were provided by their parents or legal guardians before participation.

### Procedures

Peripheral venous blood samples were collected and centrifuged to isolate serum samples, which were then stored at −20□. The titers of vaccine-induced antibodies against the EV-A71 isolate C4a were measured using a modified cytopathogenic effect (CPE) assay. Briefly, serum samples were diluted 2-fold in duplicate before use and incubated with EV-A71 isolate C4a for 2 hours. The antibody titer was calculated as the reciprocal of the lowest dilution fold of the cytopathogenicity. Samples with titers lower than 8 were assigned a value of 4 for calculation^5^. The details of CPE were depicted in **Supplementary Information Note 2**.

### Statistical analysis

The baseline characteristics of the study participants were described using medians with interquartile ranges (IQRs) for continuous variables and proportions for categorical variables. The geometric mean titers (GMTs) and geometric mean fold increase (GMFI) of EV-A71-specific antibodies, along with their 95% CIs, were calculated to describe the level and fold increase of vaccine-induced antibody titers^24^.

Antibody titers of 16 and 32 were selected as seropositive thresholds for primary and sensitive analysis, respectively^5,25^, due to the “true” seroprotective threshold having not been established yet. According to baseline titers, children were classified into positive or negative groups based on a cut-off of 8. Further, children in the positive group were defined as those with high- or low-level (i.e., <16 or ≥16) pre-vaccination titers, where the threshold was determined by the distribution of the baseline titers (**Figure S2**). Antibody titers were log-transformed using base 2 (log2) for analysis.

To explore the effect of pre-existing antibodies on vaccine-induced immunity, GMTs and GMFIs were compared based on the levels of baseline titers. To assess the differences across groups, the t-test, ANOVA test, or Mann-Whitney test was used for continuous variables, and the χ2 test or Fisher’s exact test was used for categorical variables, when appropriate. Generalized additive mixed models (GAMMs) with a spline function were constructed using a Gaussian distribution and an identity link function to assess the effects of pre-existing antibody and age at primary dose on vaccine-induced immunity. Other information, such as sex, pre-existing titers before vaccination, age at the primary dose, the date of sampling, and data source (i.e., data from an RCT study or a cohort study), was included as covariates in the GAMM model to adjust for potential confounders. The Akaike information criterion (AIC) was calculated for model selection. The details of the models are shown in **Supplementary Information Note 3**. All statistical analyses were conducted using R version 4.1.3.

## Data Availability

All data produced in the present study are available upon reasonable request to the authors

## ACKNOWLEDGMENTS

We acknowledge the support from the Shanghai Municipal Science and Technology Major Project (grant number ZD2021CY001 to H.Y.) and the Key Program of the National Natural Science Foundation of China (grant number 82130093 to H.Y.).

## AUTHOR CONTRIBUTIONS

Hongjie Yu conceptualized and supervised the study. Jiaxin Zhou designed the methodology and Xiaoyu Zhou formally analyzed the data. Xiaoyu Zhou and Jiaxin Zhou prepared the visualization presentation and original manuscript. Hongjie Yu, Junbo Chen and Nicholas C Grassly reviewed and edited the manuscript. Hongjie Yu supervised the research. All authors contributed to the manuscript and approved the final version.

## ADDTIONAL INFORMATION

Supplemental information includes details on the study design, laboratory procedures, construction of statistical models and supplemental results. It contains six figures and three tables.

## REFERENCES

1. Hong J, Liu F, Qi H, Tu W, Ward MP, Ren M, et al. Changing epidemiology of hand, foot, and mouth disease in China, 2013-2019: a population-based study. Lancet Reg Health West Pac. 2022;20:100370.

2. Mao LX, Wu B, Bao WX, Han FA, Xu L, Ge QJ, et al. Epidemiology of hand, foot, and mouth disease and genotype characterization of Enterovirus 71 in Jiangsu, China. J Clin Virol. 2010;49:100–4.

3. Tan YW, Chu JJH. Protecting the most vulnerable from hand, foot, and mouth disease. Lancet Infect Dis. 2021;21:308–9.

4. Hu YM, Wang X, Wang JZ, Wang L, Zhang YJ, Chang L, et al. Immunogenicity, safety, and lot consistency of a novel inactivated enterovirus 71 vaccine in Chinese children aged 6 to 59 months. Clin Vaccine Immunol. 2013;20:1805–11.

5. Zhu FC, Meng FY, Li JX, Li XL, Mao QY, Tao H, et al. Efficacy, safety, and immunology of an inactivated alum-adjuvant enterovirus 71 vaccine in children in China: a multicentre, randomised, double-blind, placebo-controlled, phase 3 trial. Lancet. 2013;381:2024–32.

6. Zimmermann P, Curtis N. Factors That Influence the Immune Response to Vaccination. Clin Microbiol Rev. 2019;32.

7. Gans HA, Arvin AM, Galinus J, Logan L, DeHovitz R, Maldonado Y. Deficiency of the humoral immune response to measles vaccine in infants immunized at age 6 months. Jama. 1998;280:527–32.

8. Kandeil W, Savic M, Ceregido MA, Guignard A, Kuznetsova A, Mukherjee P. Immune interference (blunting) in the context of maternal immunization with Tdap-containing vaccines: is it a class effect? Expert Rev Vaccines. 2020;19:341–52.

9. Mok DZL, Chan KR. The Effects of Pre-Existing Antibodies on Live-Attenuated Viral Vaccines. Viruses. 2020;12.

10. Saito N, Komori K, Suzuki M, Morimoto K, Kishikawa T, Yasaka T, et al. Negative impact of prior influenza vaccination on current influenza vaccination among people infected and not infected in prior season: A test-negative case-control study in Japan. Vaccine. 2017;35:687–93.

11. Tong Y, Zhang X, Chen J, Chen W, Wang Z, Li Q, et al. Immunogenicity and safety of an enterovirus 71 vaccine in children aged 36-71 months: A double-blind, randomised, similar vaccine-controlled, non-inferiority phase III trial. EClinicalMedicine. 2022;52:101596.

12. Zhang L, Gao F, Zeng G, Yang H, Zhu T, Yang S, et al. Immunogenicity and Safety of Inactivated Enterovirus 71 Vaccine in Children Aged 36-71 Months: A Double-Blind, Randomized, Controlled, Non-inferiority Phase III Trial. J Pediatric Infect Dis Soc. 2021;10:440–7.

13. Hodgson D, Sánchez-Ovando S, Carolan L, Liu Y, Hadiprodjo AJ, Fox A, et al. Quantifying the impact of pre-vaccination titre and vaccination history on influenza vaccine immunogenicity. Vaccine. 2025;44:126579.

14. van der Staak M, Ten Hulscher HI, Nicolaie AM, Smits GP, de Swart RL, de Wit J, et al. Long-term dynamics of measles virus-specific neutralizing antibodies in children vaccinated before 12 months of age. Clin Infect Dis. 2024.

15. Sasaki S, He XS, Holmes TH, Dekker CL, Kemble GW, Arvin AM, et al. Influence of prior influenza vaccination on antibody and B-cell responses. PLoS One. 2008;3:e2975.

16. Halsey N, Galazka A. The efficacy of DPT and oral poliomyelitis immunization schedules initiated from birth to 12 weeks of age. Bull World Health Organ. 1985;63:1151–69.

17. di SAPA. Simultaneous immunization of newborn infants against diphtheria, tetanus, and pertussis; production of antibodies and duration of antibody levels in an eastern metropolitan area. Am J Public Health Nations Health. 1950;40:674–80.

18. Nair N, Gans H, Lew-Yasukawa L, Long-Wagar AC, Arvin A, Griffin DE. Age-dependent differences in IgG isotype and avidity induced by measles vaccine received during the first year of life. J Infect Dis. 2007;196:1339–45.

19. Huang ML, Chiang PS, Chia MY, Luo ST, Chang LY, Lin TY, et al. Cross-reactive neutralizing antibody responses to enterovirus 71 infections in young children: implications for vaccine development. PLoS Negl Trop Dis. 2013;7:e2067.

20. Wei X, Yang J, Gao L, Wang L, Liao Q, Qiu Q, et al. The transfer and decay of maternal antibodies against enterovirus A71, and dynamics of antibodies due to later natural infections in Chinese infants: a longitudinal, paired mother-neonate cohort study. Lancet Infect Dis. 2021;21:418–26.

21. Nic Lochlainn LM, de Gier B, van der Maas N, Strebel PM, Goodman T, van Binnendijk RS, et al. Immunogenicity, effectiveness, and safety of measles vaccination in infants younger than 9 months: a systematic review and meta-analysis. Lancet Infect Dis. 2019;19:1235–45.

22. Li JX, Song YF, Wang L, Zhang XF, Hu YS, Hu YM, et al. Two-year efficacy and immunogenicity of Sinovac Enterovirus 71 vaccine against hand, foot and mouth disease in children. Expert Rev Vaccines. 2016;15:129–37.

23. Li R, Liu L, Mo Z, Wang X, Xia J, Liang Z, et al. An inactivated enterovirus 71 vaccine in healthy children. N Engl J Med. 2014;370:829–37.

24. Nauta J. Statistics in Clinical Vaccine Trials. Heidelberg: Springer Berlin; 2011.

25. Chou AH, Liu CC, Chang JY, Jiang R, Hsieh YC, Tsao A, et al. Formalin-inactivated EV71 vaccine candidate induced cross-neutralizing antibody against subgenotypes B1, B4, B5 and C4A in adult volunteers. PLoS One. 2013;8:e79783.

