## Supplementary for "Pre-existing antibodies and age shape the immune response following EV-A71 vaccine: a prospective serological study among the Chinese pediatric population"

**Supplementary Information**

### Note 1.  **Details of the study design**

#### 1.1 **Phase 3 clinical trial**

(1) Inclusion and exclusion criteria

Healthy children aged 6 to 35 months of age were recruited from Sichuan Province between September 2019 and April 2020 to assess the equivalence of the immunity responses of three batches of inactivated EV-A71 vaccines as well as the immunogenicity, safety, and tolerability of the vaccines among healthy children. A total of 720 children were stratified into three age strata of 6-11, 12-23 and 24-35 months of age, with 240 children in each stratum. Exclusion criteria for the study included a history of EV-A71 vaccination or HFMD, a history of vaccine allergies, or the presence of any acute or severe underlying diseases. Participants were also excluded if they had received immunoglobulin or a blood transfusion within three months of the first injection, or if they had received any other inactivated vaccine within one week or a live vaccine within two weeks of enrollment. Finally, any other health conditions that deemed a child unsuitable for participation, as determined by the study's trained project staff, also resulted in exclusion.

(2) Randomization and administration

After physical examination, children were randomized into three groups to receive one of the three batches of inactivated EV-A71 vaccine. All children were scheduled to receive two doses of the vaccine to complete the full vaccination course (one primary and one booster dose), with an interval of 30 days between two doses. Three batches of the experimental vaccines (batch number: 201809046, 201809050, and 201810054) were supplied by SINOVAC BIOTECH Co., Ltd. (Beijing, China), which were made available in 0.5 mL per dose for intramuscular injection in the deltoid muscle.

(3) Blood sampling

Paired blood samples were collected from all participants before the first dose (day 0) and 30 days after full vaccination (day 60). Additionally, a subset of 270 children was selected through age stratification (i.e., 6-11 months of age, 12-23 months of age, and 24-35 months of age) to provide additional blood samples at five timepoints: 7 day, 14 days and, 30 days after the first dose, 7 days and 14 days after the second dose. At each time point, blood samples were collected from 54 children (**Figure 1, Figure S1**).

#### 1.2 **Cohort study**

(1) Inclusion and exclusion criteria

A total of 9 570 children were recruited in the cohort study during April 2019 and March 2020 in Xiantao and Songzi City, Hubei Province. Exclusion criteria for this study include a history of HFMD, unclear baseline information, history of EV-A71 vaccine before enrollment, and any other health conditions that were not suitable for participation.

(2) Administration and blood sampling

During follow-up period, 4 512 (47.1%) children received primary and booster dose of EV-A71 vaccine and have paired serum samples before and after full immunization. Children who were not sampled at baseline or post-vaccination were also excluded for data analysis (**Figure 1, Figure S1**).

#### 1.3 **The details of blood sampling time**

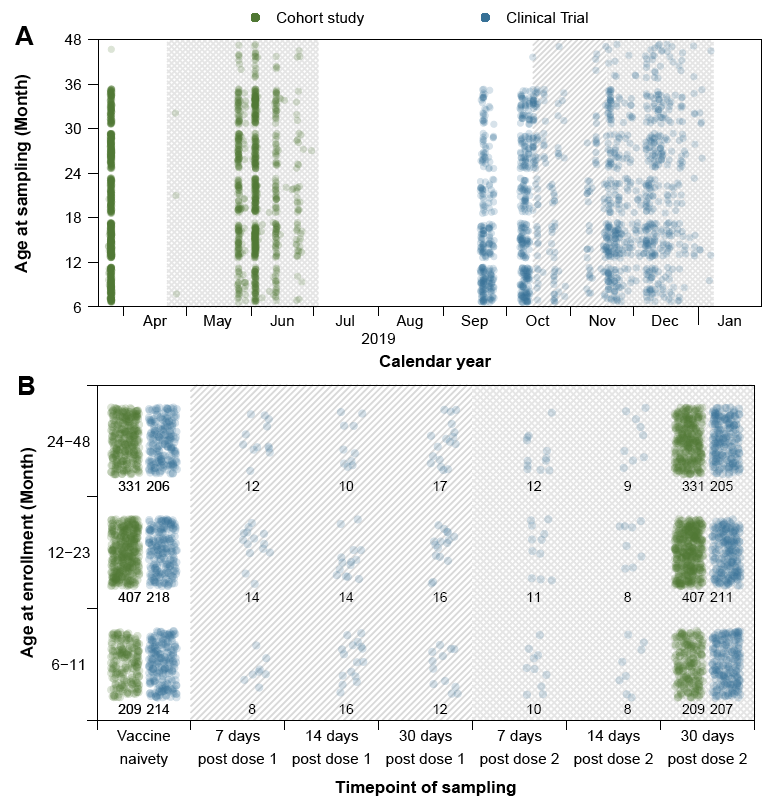

##### **Figure S1. Detailed timepoint of blood sampling and sample size of participants from cohort study and clinical trial.**

Panel (A) shows the calendar time of sampling. Panel (B) shows the timepoint of sampling relative to the date of vaccination. The numbers in panel (B) represent the sample size by age group at each timepoint. Note that green and blue represent the different sources of data. Each dot represents sample at individual level at a given timepoint. The white, hatched, and cross-hatched areas indicate the different vaccine status (i.e., vaccine naïvety, after first dose, and after second dose), respectively.

### Note 2.  **Laboratory procedures**

The peripheral venous blood samples were collected and centrifuged to isolate serum samples before stored at -20℃. The levels of neutralizing antibodies against EV-A71 isolate C4a were measured by the modified cytopathogenic effect (CPE) assay. The serum samples were 1:8 diluted before use and were inactivated at 56℃ for 30 min. Each well of the 96-well plate was added with 50 μL serum samples at 2-fold dilution and each sample were tested in duplicate. The maximum dilution factor was 2^14^ and 2^15^ in the RCT and the cohort study, respectively. Cell controls were also set during the experiment. After adding diluted EV-A71 with a 50% cell culture infective dose (CCID50) of 100/0.05mL, the 96-well plates were incubated at 37℃ for 2 hours. Human rhabdomyosarcoma cells (1.5-2*10^5^) were then added to the serum-virus suspensions with a volume of 100μL per well. Cytopathogenic results were obtained after incubation for 5-7 days and the titer of neutralization antibodies in serum samples were calculated as the reciprocal of the serum dilution of the cytopathogenicity. For samples in the RCT study with antibody titers exceeding 2^14^, we assigned a value of 2^15^ for subsequent quantitative analysis.

### Note 3.  **Construction of generalized additive models**

#### 3.1 **Model for assessing the effect of pre-existing antibodies and the age at primary dose among children with pre-existing antibody**

The model description has been provided below:

$$g\left( E\left[ y_{i} \right] \right)=\alpha+\beta_{1}s\left( {log\_titer0}_{i} \right)+\beta_{2}s\left( {age\_dose1}_{i} \right)+\beta_{3}{sex}_{i}+\beta_{4}s\left( {sample\_date}_{i} \right)+\beta_{5}{group}_{i}+\mu_{0}$$

where $g()$ is the identity link function and $y_{i}$ is log-transformed vaccine-induced antibody titers or the fold increase for the $i$th child post full vaccination. $\alpha$ is the intercept. $s\left( {log\_titer0}_{i} \right)$, $s\left( {age\_dose1}_{i} \right)$ and $s\left( {sample\_date}_{i} \right)$ are the smooth functions used to characterize the effect of log-transformed antibody titer prior to vaccination, age at the first dose, and the date of sampling, respectively. ${sex}_{i}$ and ${group}_{i}$ refer to the sex of participants and the data source, respectively. $\beta_{1}$ to $\beta_{5}$ represent the coefficients. $\mu_{0}$ denotes the random effect of children following repeated measurements of antibody concentration. In the sensitive analysis, children without pre-existing antibody were also included for modeling. Furthermore, we also explored the effect of pre-existing antibody across doses.

To further assess the robustness of the age effect, we further constructed 2 alternative models as given below:

*Alternative Model 1*: Linear regression with age and all other covariates included as linear terms.

*Alternative Model 2*: GAMM with age included as a linear component, while maintaining the original form of other variables as in the main analysis model.

The goodness of fit between primary model and two alternative models were shown in **Table S1**, showing a relative superior goodness-of-fit for our primary GAMM approach.

##### **Table S1. Summary of statistics of goodness of fit between primary model and alternative models.**

| **Statistics** | **R^2^** | **AIC** |
| --- | --- | --- |
| Primary model | 0.167 | 790.272 |
| Alternative Model 1 | 0.157 | 792.944 |
| Alternative Model 2 | 0.152 | 791.356 |

#### 3.2 **Model for assessing the effect of the age at primary dose among children without pre-existing antibody**

The model descriptions were provided below:

$$g\left( E\left[ y_{i} \right] \right)=\alpha+\beta_{1}s\left( {age\_dose1}_{i} \right)+\beta_{2}{sex}_{i}+\beta_{3}s\left( {sample\_date}_{i} \right)+\beta_{4}{group}_{i}+\mu_{0}$$

where $g()$ is the identity link function and $y_{i}$ is log-transformed antibody titer or the fold increase for the $i$th child post full vaccination. $\alpha$ is the intercept. $s\left( {age\_dose1}_{i} \right)$ and $s\left( {sample\_date}_{i} \right)$ are the smooth functions used to characterize the effect of age at the first dose and the date of sampling, respectively. ${sex}_{i}$ and ${group}_{i}$ refer to the sex of participants and the data source, respectively. $\beta_{1}$ to $\beta_{4}$ represent the coefficients. $\mu_{0}$ denotes the random effect of children following repeated measurements of antibody concentration.

Similarly, 2 alternative models were constructed as given below to assess the appropriateness of models we chose for analysis, and better goodness of fit was observed in the primary GAMM (**Table S2**).

*Alternative Model 1*: Linear regression with age and all other covariates included as linear terms.

*Alternative Model 2*: GAMM with age included as a linear component, while maintaining the original form of other variables as in the main analysis model.

##### **Table S2. Summary of statistics of goodness of fit between primary model and alternative models.**

| **Statistics** | **R^2^** | **AIC** |
| --- | --- | --- |
| Primary model | 0.110 | 801.048 |
| Alternative Model 1 | 0.086 | 807.123 |
| Alternative Model 2 | 0.060 | 807.123 |

### Note 4.  **Characteristics of participants**

#### 4.1 **Baseline characteristics comparison between selected and non-selected children**

We compared the basic characteristics of children selected and not selected for analysis in this study, and observed a higher proportion of boys among non-selected children (***p*** = 0. 0017, **Table S3**). The age at baseline is similar between groups (***p*** = 0.1088).

##### **Table S3. Baseline characteristics comparison between participants and non-participants.**

| **Characteristics** | **Selected children**  **(N = 1 585)** | **Non-selected children**  **(N = 8 705)** | **All**  **(N = 10 290)** | ***p* value** |
| --- | --- | --- | --- | --- |
| **Sex** |  |  |  |  |
| Boys | 796 (50.22) | 4 747 (54.53) | 5 543 (53.87) | **0.0017** |
| Girls | 789 (49.78) | 3 958 (45.47) | 4 747 (46.13) |  |
| **Age at baseline (month) ^a^** | |  |  |  |
| Median (IQR) | 17.5 (11.3-26.9) | 20.3 (12.7-29.1) | 19.9 (12.4-28.8) |  |
| 6-11 | 423 (26.69) | 1 938 (22.27) | 2 361 (22.95) | 0.1088 |
| 12-23 | 625 (39.43) | 3 312 (38.06) | 3 937 (38.27) |  |
| 24-35 | 536 (33.812) | 2 691 (30.92) | 3 227 (31.37) |  |

Data are expressed as n (%) unless otherwise specified.

^a^ Numbers in each subgroup may not sum up to 1 585, 8 705 and 10 290 due to missing information.

### Note 5.  **Description of the levels of pre-existing antibody titer**

#### 5.1 **The distribution of pre-exiting antibody titer**

The distribution of antibody titers before vaccination was depicted in **Figure S2**. According to the distribution of pre-existing antibody, 16 was chosen as the threshold to further group children with pre-existing antibodies into “children with high level of pre-existing antibodies” and “children with low level of pre-existing antibodies”.

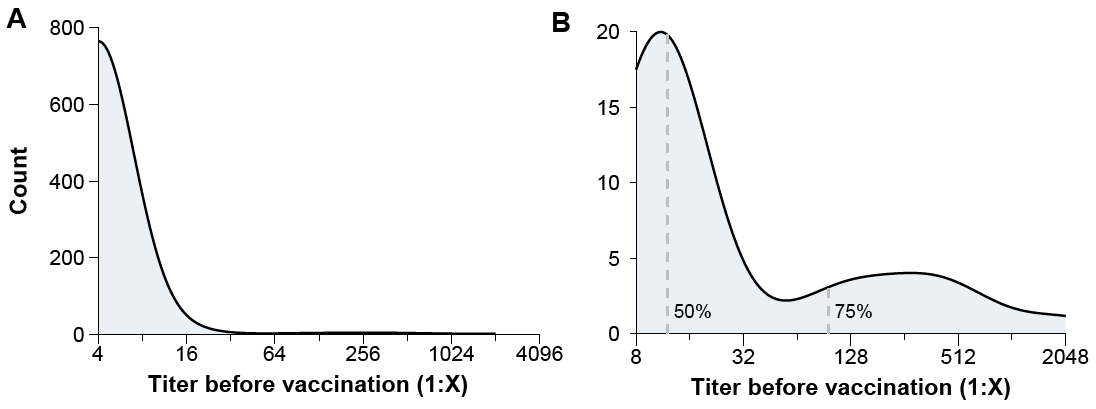

##### **Figure S2. Distribution of pre-existing antibodies.**

Panel (A) and (B) show the distribution of pre-existing antibody before vaccination among all participants and among participants with pre-existing antibody titer, respectively.

#### 5.2 **The individual antibody titers and GMTs across timepoints**

According to the dynamics of individual antibody level and GMT, we can observe that children with pre-existing antibody mounted a higher level of vaccine-induced antibody titer and a lower magnitude of antibody increase, compared to children without pre-existing antibody (**Figure S3**).

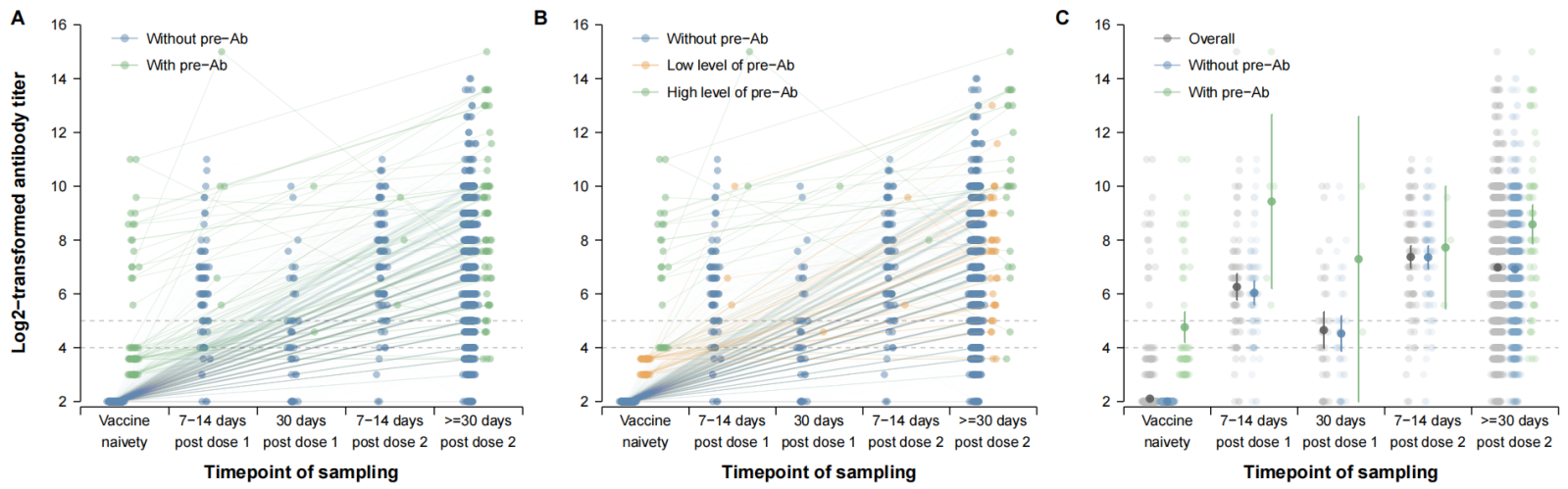

##### **Figure S3. Antibody titers at different sampling timepoints, by levels of pre-existing antibody.**

Panel (A) and (B) show the individual antibody trajectories and panel (C) shows the GMTs at different sampling timepoints. Participants are divided into 2 groups (i.e., with or without pre-existing antibody before vaccination) in panel (A) and (C), while divided into 3 groups (i.e., with high levels of pre-existing antibody, low levels of pre-existing antibody, or without pre-existing antibody) in panel (B). Note that the dashed grey lines in the three panels denote the selected seropositive thresholds (i.e., log-transformed titers of 4 and 5).

### Note 6.  **Potential effect of pre-existing antibody titer across doses**

#### 6.1 **Immunity response across doses, by level of pre-existing antibody**

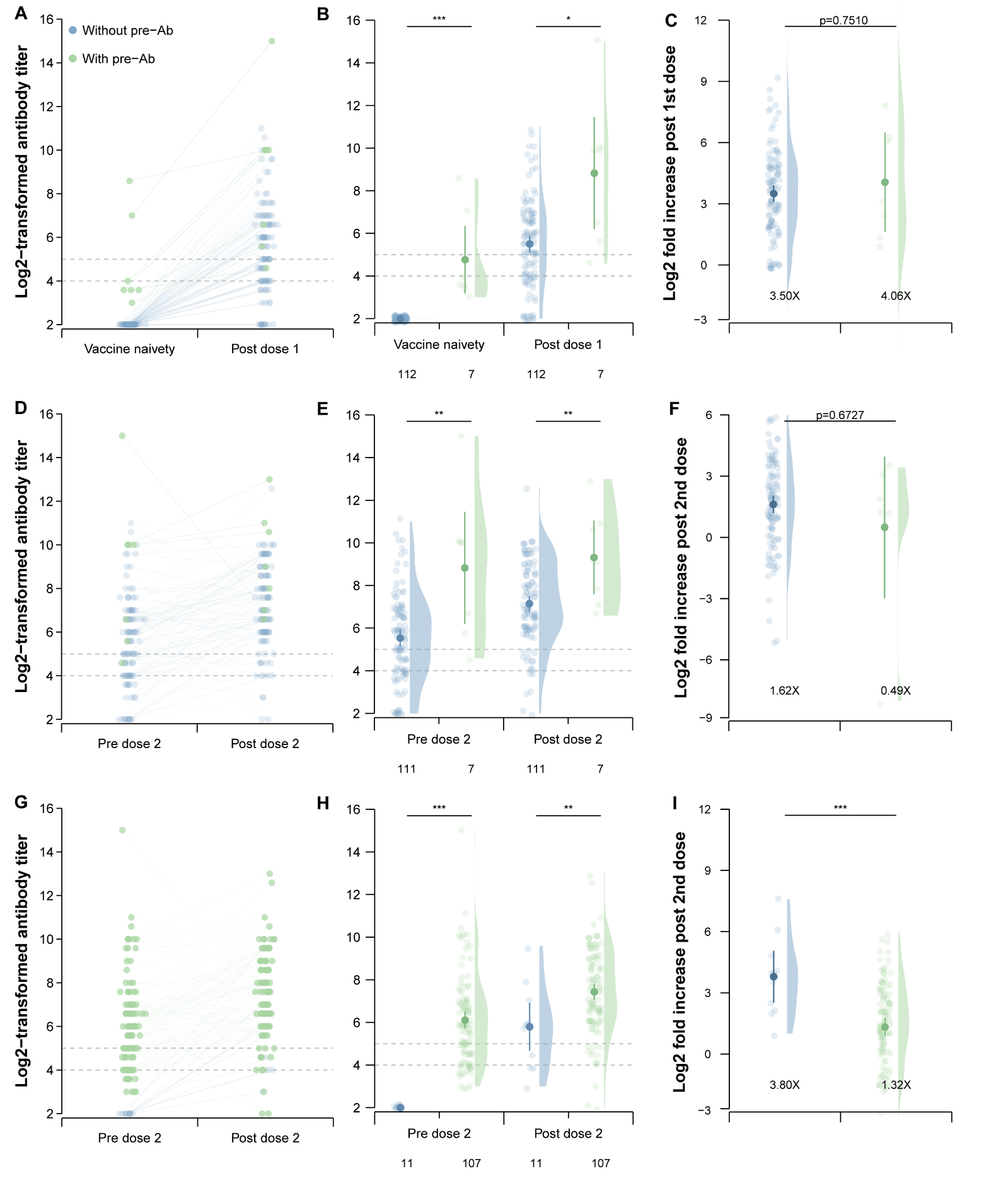

##### **Figure S4. Immunity response across doses, by level of pre-existing antibody.**

Panel (A)-(C) show the immune response from vaccine naïvety to 7-14 days after receiving the first dose, by the level of antibody titers before dose 1. Panel (D)-(F) show the immune response from before to after receiving the second dose, by the level of antibody titers before dose 1. Panel (G)-(I) show the immune response from before to after receiving the second dose, by the level of antibody titers before dose 2. Individual antibody trajectories across doses are shown in panel (A), (D) and (G). Individual antibody levels and GMTs pre- and post-vaccination are shown in panel (B), (E) and (H), and the figures at the bottom of 3 panels represent sample sizes of each group. Log2-transformed fold increase of antibody titers induced by each dose are shown in panel (C), (F) and (I), and the geometric means of fold increase were marked at the bottom. “*”, “**” and “***” represent the *p*-value of statistic test is <0.05, <0.01 and <0.001. The dashed grey lines denote the selected seropositive thresholds (i.e., log-transformed titers of 4 and 5).

#### 6.2 **Analyzing the effect of pre-existing antibodies using GAMMs, among children with and without pre-existing titers**

After including the data from children with undetectable pre-existing antibodies, we consistently found the antibody titers following two doses of vaccination exceeding the pre-defined seropositivity threshold, irrespective of pre-vaccination titers. Vaccine-induced antibody titer increased with log-transformed pre-vaccination titers and reached their peak of 9.07 (95% CI: 8.10, 9.15) when the pre-existing titer was 10.62 (**Figure S5**), when the effects of potential confounders were controlled. However, the fold increase in antibody titers declined with higher pre-existing antibody levels with a lowest level of 1.89 (95% CI: -0.64, 4.41) when antibody titers before vaccination was 12.00 (**Figure S5**).

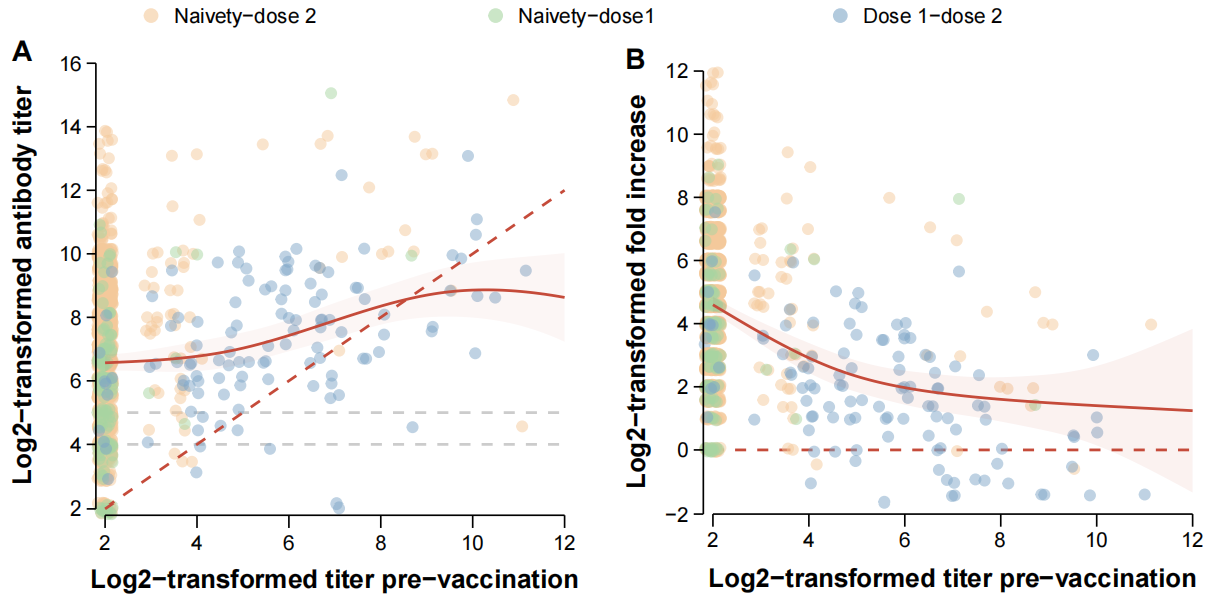

##### **Figure S5. Fitted vaccine-induced antibody titers and fold increase using GAMMs, among children with and without pre-existing antibody.**

Panel (A) shows the scatters and fitted line of vaccine-induced antibody titers by the levels of pre-existing titer. Panel (B) shows the scatters and fitted line of fold increase post-vaccination by the levels of pre-existing titer. Note that the solid lines and shadows indicate the fitted mean and 95% CI band. The brown dashed lines in two panels indicate the reference line. The grey dashed lines denote the selected seropositive thresholds (i.e., log-transformed titers of 4 and 5).

#### 6.3 **Analyzing the effect of pre-existing antibodies across doses using GAMMs, among children with pre-existing titers**

To further validate the robustness of the findings, we stratified the data by doses and fitted GAMMs with each subset. The results were consistent with those of the primary analysis, indicating the results were subtly affected by the dose of vaccination.

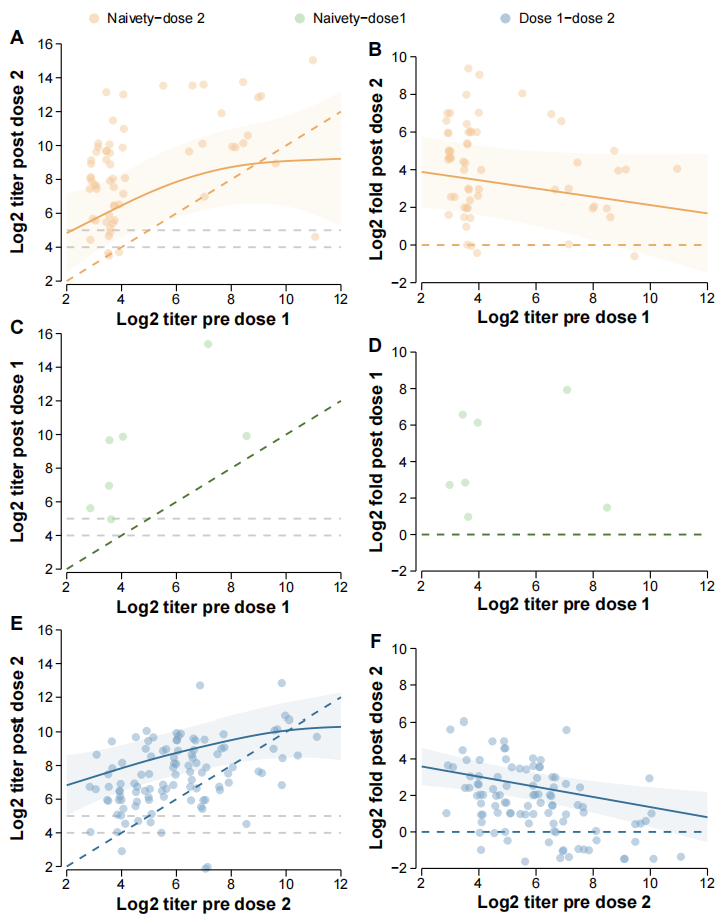

##### **Figure S6. Fitted post-vaccination antibody titers and fold increase across doses.**

Panel (A)-(B) show the scatters and fitted line of antibody titers and fold increase post 2 doses of vaccination, respectively. Panel (C)-(D) show the scatters of dose-1-induced antibody titers and fold increase, respectively. No fitted line is shown in panel (C)-(D) due to the limitation of sample size. Panel (E)-(F) show the scatters and fitted line of dose-2-induced antibody titers and fold increase, respectively. The solid lines and shadows indicate the fitted mean and 95% CI band. The orange, green, and blue dashed lines indicate the reference line. The grey dashed lines denote the selected seropositive thresholds (i.e., log-transformed titers of 4 and 5).
